# Race/Ethnic Disparities in Mild Cognitive Impairment and Dementia: The Northern Manhattan Study

**DOI:** 10.1101/2020.11.07.20210872

**Authors:** Clinton B. Wright, Janet T. DeRosa, Michelle P. Moon, Kevin Strobino, Charles DeCarli, Ying Kuen Cheung, Stephanie Assuras, Bonnie Levin, Yaakov Stern, Xiaoyan Sun, Tatjana Rundek, Mitchell S.V. Elkind, Ralph L. Sacco

**Affiliations:** National Institute of Neurological Disorders and Stroke, national Institutes of Health, Bethesda, MD; Department of Neurology, Vagelos College of Physicians & Surgeons, Columbia University, New York, NY; Department of Neurology, University of California at Davis, Davis, CA; Department of Biostatistics, Mailman School of Public Health, Columbia University, New York, NY; Evelyn F McKnight Brain Institute, University of Miami Miller School of Medicine, Miami, FL; Department of Neurology, University of Miami Miller School of Medicine, Miami, FL

**Keywords:** dementia, mild cognitive impairment, cohort studies, Hispanic American, African American

## Abstract

**OBJECTIVE:** Estimate the prevalence of mild cognitive impairment (MCI) and probable dementia in the racially and ethnically diverse community-based Northern Manhattan Study cohort and examine sociodemographic, vascular risk factor, and brain imaging correlates.

**METHODS:** Cases of MCI and probable dementia were adjudicated by a team of neuropsychologists and neurologists and prevalence was estimated across race/ethnic groups. Ordinal proportional odds models were used to estimate race/ethnic differences in prevalence rates for MCI or probable dementia adjusting for sociodemographic variables (model 1), model 1 plus potentially modifiable vascular risk factors (model 2), and model 1 plus structural imaging markers of brain integrity (model 3).

**RESULTS:** There were 989 participants with cognitive outcome determinations (mean age 69 ± 9 years; 68% Hispanic, 16% Black, 14% White; 62% women; mean (±SD) follow-up five (±0.6) years). Prevalence rates for MCI (20%) and probable dementia (5%) were significantly different by race/ethnicity even after accounting for age and education difference across race-ethnic groups; Hispanic and Black participants had greater prevalence rates than Whites. Adjusting for sociodemographic and brain imaging factors explained the most variance in the race/ethnicity associations. White matter hyperintensity burden explained much of the disparity between Black and White, but not between Hispanic and White, participants.

**CONCLUSIONS:** In this diverse community-based cohort, white matter hyperintensity burden partially explained disparities in MCI and dementia prevalence in Black but not Hispanic participants compared to Whites. Longer follow-up and incidence data are needed to further clarify these relationships.

## INTRODUCTION

Variability in dementia rates across racial and ethnic groups has been estimated at 60%, underscoring the importance of work to understand ethnoracial disparities.^1^ Improving our understanding and reducing rates of cognitive impairment and dementia disparities is a major goal of the National Alzheimer’s Plan (NAPA). Understanding the factors underlying these disparities, including the importance of modifiable risk factors and susceptibility to pathological processes and resistance to them, is a priority to appropriately target interventional strategies.

Research in diverse cohorts plays an important role in clarifying race/ethnic differences, and disparities in the risk of mild cognitive impairment (MCI) and dementia have been reported for Black people in multiple studies over the past decades,^1-4^ but data on Hispanics is more limited. Diverse cohorts that include multiple ethnic and racial groups with deep phenotyping of key behavioral and modifiable risk factor exposures, intermediate markers of brain integrity, and cognitive outcomes are needed to improve our understanding of disparities, especially in vascular cognitive impairment and dementia (VCID) risk. White matter hyperintensities, subclinical brain infarcts, and brain atrophy measures detected by MRI can be particularly helpful in this regard. We examined ethnoracial disparities in prevalence rates of mild cognitive impairment (MCI) and dementia in a diverse population-based cohort of Hispanic, Black, and White people living in the Northern Manhattan community of New York City.

## MATERIAL AND METHODS

### Study Population

The NOMAS sample enrolled adults aged 40 and older (range 40-94) who had resided in Northern Manhattan for more than three months at the time of recruitment and had never been diagnosed with a stroke^18^. Participants were recruited between 1993-2001 using random-digit dialing to participate in a baseline interview and assessment (enrollment response rate was 75%), with ongoing annual telephone and in-person follow-up (N=3,298; loss to follow up <5%). In 2003 the MRI sub-study began recruitment during annual telephone follow-up and included NOMAS participants who were clinically stroke-free, age 50 and older, and had no contraindications to MRI. An additional 199 household members were recruited to yield a final sample size of 1,290 by 2008. The study is approved by the IRBs of Columbia University Medical Center and the University of Miami School of Medicine and all subjects provided written informed consent.

### Baseline Evaluation

Participants had a thorough evaluation of vascular risk factors and medical history at the time of enrollment, including a physical/neurological examination by study physicians. Race/ethnicity was defined by self-identification using a series of questions modeled after the US census and conforming to standard definitions outlined by Directive 15. Years of educational attainment, including degree achieved, were self-reported. Standardized questions were adapted from the CDC Behavioral Risk Factor Surveillance System regarding history of hypertension, diabetes mellitus, smoking, and cardiovascular conditions, including congestive heart failure, angina, coronary artery disease, atrial fibrillation, valvular heart disease, and peripheral vascular disease^20,21^. Data on medication use was collected. Smoking was categorized as current (within the past year), former, or never smoker of cigarettes, cigars, or pipes. Leisure-time physical activity was assessed by self-report using a questionnaire adapted from the National Health Interview Survey and moderate to heavy physical activity was defined as participation in at least one of several rigorous physical activities in a typical 14-day period.^22^ Moderate alcohol use was defined as current drinking of one drink per month up to two drinks per day. Fasting blood specimens were analyzed at the Core Laboratory of the Irving Center for Clinical Research to determine blood glucose and lipid levels, including total cholesterol and HDL. Waist and hip circumferences were measured in inches with a flexible tape measure while participants were standing and wearing no heavy outer garments. Waist circumference (WC) was measured at the level of the umbilicus, and hip circumference was measured at the level of the bilateral greater trochanters, as previously described.^5^ Systolic and diastolic blood pressures were calculated by averaging two measurements (before and after the physical examination) from the right brachial artery after a 10-minute rest in a seated position (Dinamap Pro100, Critikon Inc).

### Brain MRI

Imaging was performed on a 1.5-Tesla MRI system (Philips Medical Systems, Best, The Netherlands) at the Columbia University Hatch Research Center. To segment white matter hyperintensities, semiautomated measurements of pixel distributions using mathematical modeling of pixel-intensity histograms for cerebrospinal fluid and brain white and gray matter were used to identify the optimal pixel-intensity threshold to distinguish cerebrospinal fluid from brain matter, using a custom-designed image analysis package (QUANTA 6.2 using a Sun Microsystems Ultra 5 workstation).^6^ For subclinical brain infarct readings, methods to identify and classify MRI-defined subclinical infarcts (SBI) have been published.^7^ Two independent raters used a superimposed image of the subtraction, fluid attenuated inversion recovery (FLAIR), proton density, and T2-weighted images at 3× magnified view for interpretation of lesion characteristics. Agreement among raters has been good (published kappa values, 0.73–0.90).^8^ Processing of MRI scans to calculate total intracranial volumes (ICV), total cerebral volumes, and white matter hyperintensity volumes (WMHV), has been previously described.^9^ To correct for head size, WMHV was calculated as percent total ICV and log-transformed to a normal distribution (log-WMHV). Proportion of total cerebral volume to total ICV was examined as brain parenchymal fraction (BPF). Subclinical infarcts (SBI) were rated as present or absent.

### Cognitive and Functional Assessment

Participants in the MRI sub-study were recruited between 2003 to 2008 and assessed in-person (defined as visit 1) and were interviewed by a trained research assistant, who administered structured questionnaires and a neuropsychological (NP) battery. All tests were administered in English or Spanish based on participant preference. The Mini Mental State Examination and Cognitive Failures Questionnaire were administered at the time of the NP battery in a designated quiet room. Most testing was on the day of MRI.^10, 11^ Episodic memory was measured using three sub-scores derived from a 12-word five trial list-learning task: list learning total score, delayed recall score, and delayed recognition score. Executive function was assessed with two sub-scores: the difference in time to complete the Color Trails test Form 1 and Form 2, and the sum of the Odd-Man-Out subtests 2 and 4.^12^ Processing speed was assessed with the Grooved Pegboard task non-dominant hand time and the Color Trails test Form 1.^13, 14^ Working memory was assessed with the Digit Ordering and Letter Number Sequencing tests.^15, 16^ Semantic memory was measured using three tests: picture naming (modified Boston Naming), category fluency (Animal Naming) and phonemic fluency (C, F, L in English speakers and P, S, V in Spanish speakers).^17, 18^ The Visual-Motor Integration test and the Symbol Digit Modalities tests were not part of the domains but were also available for review.^13, 19^ At the initial visit we estimated premorbid intelligence and literacy with the Peabody Picture Vocabulary Test (Test de Vocabulario en Imagenes Peabody for Spanish speakers), the Wide Range Achievement Test (English speakers) and the Word Accentuation Test (Spanish speakers).^20-22^ Depressive symptoms were quantified with the Center for Epidemiological Studies Depression scale (CES-D), a 20-item scale assessing depressive affect, somatic complaints, positive affect, and interpersonal relations.^23^ Research assistants interviewed participants to assess cognitive functional status.

A mean of 5.0±0.6 years after the initial neuropsychological assessment, a second in-person assessment was conducted between 2008 and 2015 (visit 2). At the second visit, the neuropsychological battery was repeated with the addition of the Letter Number Sequencing and Symbol Digit Modalities tests, and the Informant Questionnaire of Cognitive Decline in the Elderly (IQCODE) was administered to a family member or friend of the participant as close in time to the neuropsychological assessment as possible.^25^ The relationship of the informant to the participant was documented and aided adjudication.

### Ascertainment of mild cognitive impairment and probable dementia

In 2015 NOMAS entered its fifth consecutive grant cycle and for the first time was funded to ascertain MCI and dementia status. A team of neuropsychologists and neurologists with dementia expertise used established criteria for case ascertainment.^26-28^ Each pair of adjudicators reviewed data from both visit 1 and visit 2 and completed a visit 2 grading form documenting cognitive status as normal, MCI (with notation of amnestic and non-amnestic sub-types), probable dementia, other psychiatric disorder, or unable to classify.

To assess cognitive performance, we used individual neuropsychological test scores and cognitive domain-specific scores. Cognitive domains (episodic memory, semantic memory, processing speed and executive function) were created based on an exploratory factor analysis and prior findings. Because NOMAS participants have low educational attainment, literacy, and socio-economic status, and there are limited established norms that consider these factors, we constructed NOMAS-specific norms using neuropsychological test scores from visit 1. The normative values for each NP test were calculated from age (50-60, 61-70, 71-80 and >80 years old) and education (0-6, 7-12 and >12 years of education) group-specific means and standard deviations. Each NP test score was standardized against its normative values, and each cognitive domain Z score was obtained by taking the average of the standardized NP test scores for the available component tests in a given domain.

Adjudicators accessed a Redcap web portal that displayed demographic information, visit eyesight and hearing status, neuropsychological domain and literacy Z-scores, dementia rating, MMSE and CES-D scores, most recent medication list and prior history of antidepressant and other psychiatric medications, psychiatric history, cognitive failures questionnaire, IQCODE score (cutoff for probable dementia >3.6), and stroke history between visits (if applicable).^11^ Downloadable case-specific test forms were available for review to aid in assessments. Adjudicators were randomly assigned cases blinded to each other’s ratings based on an algorithm that identified participants with possible cognitive impairment. We derived an algorithm to segregate cases who had neuropsychological testing at Visit 2 into one of two pools: 1) those requiring adjudication, and 2) those rated as cognitively normal based on an algorithm. The criteria for consensus adjudication were:

- Missing at least one of the 11 NP battery <u>tests</u>; or
- Two or more age and education normalized NP <u>test</u> Z-scores <-1.5; or
- One or more <u>domain</u> Z-scores <-1.5.

All available information pertaining to medical history, informant reports, self-rated questionnaires, educational attainment, literacy, notes from the participant interview at the visit, and other data were used for case ascertainment, but rule-based criteria helped guide cognitive status determination as follows:

- Normal: no cognitive <u>domain</u> Z-scores below -1.5, no evidence of dementia on the participant interview or IQCODE (score < 3.6).
- MCI: any <u>domain</u> Z-score below -1.5 <u>and</u> no evidence of dementia on the IQCODE (score < 3.6).
- Probable dementia: any <u>domain</u> Z-score below -1.5 <u>and</u> an IQCODE score 3.6 or greater.

Discordant ratings were resolved by consensus of the neuropsychologist/neurologist adjudication pair, and disagreements were resolved through interdisciplinary Dementia Consensus Committee (DCC) reviews.

### Statistical Analysis

Participant characteristics were compared across race/ethnicities. Characteristics between participants who only attended visit 1 and those who returned for visit 2 were also compared. We used ordinal proportional odds models to examine race/ethnic differences in prevalence using a three-level ordinal dependent variable of normal, MCI, and probable dementia, and difference of association across the three ethnic groups collectively was tested using a likelihood ratio test with 2 degrees of freedom (LRT 2 df). Less than 2% of the sample self-identified as other race (Asian or Pacific Islander, American Indian or Alaskan Native) and were excluded from this analysis. Hispanic and non-Hispanic Black (referred to as Black) were entered into the model (reference=White). Model 1 was adjusted for age, sex, years of educational attainment, medical insurance status (proxy for income), and literacy; model 2 was adjusted for variables in model 1 plus systolic and diastolic blood pressure, antihypertensive medication use, diabetes mellitus, smoking status, moderate alcohol use, physical activity, low and high density lipoprotein levels, and waist-to-hip ratio; model 3 was adjusted for variables in model 1 plus brain parenchymal fraction, log-WMHV, and SBI status. We tested for potential effect modifiers of race and ethnicity by entering multiplicative terms for significant covariates into the model. We performed a sensitivity analysis among participants with normal cognition at visit 1 (MMSE >26) to limit confounding by pre-morbid cognitive problems. Analyses were conducted using SAS 9.4 (SAS Institute, Cary, NC).

## RESULTS

Characteristics of the study sample are shown in Table 1 (mean age=69, IQR 63-74; 68% Hispanic, 16% Black, 14% White, 2% other race). Compared to participants who only attended visit 1, those who returned for visit 2 (flow across visits shown in the Figure) were younger (OR=0.96, 95%CI=0.93 to 0.99), more likely to be women (OR=1.6, 95% CI 1.0 to 2.3), had lower systolic (OR=0.7, 95% CI=0.6 to 0.9) and higher diastolic (OR=1.3, 95% CI=1.1 to 1.6) blood pressures (BP), and had better global cognitive domain scores at visit 1 (OR=1.8 per SD, 95% CI=1.4 to 2.5; Supplementary Table 1).

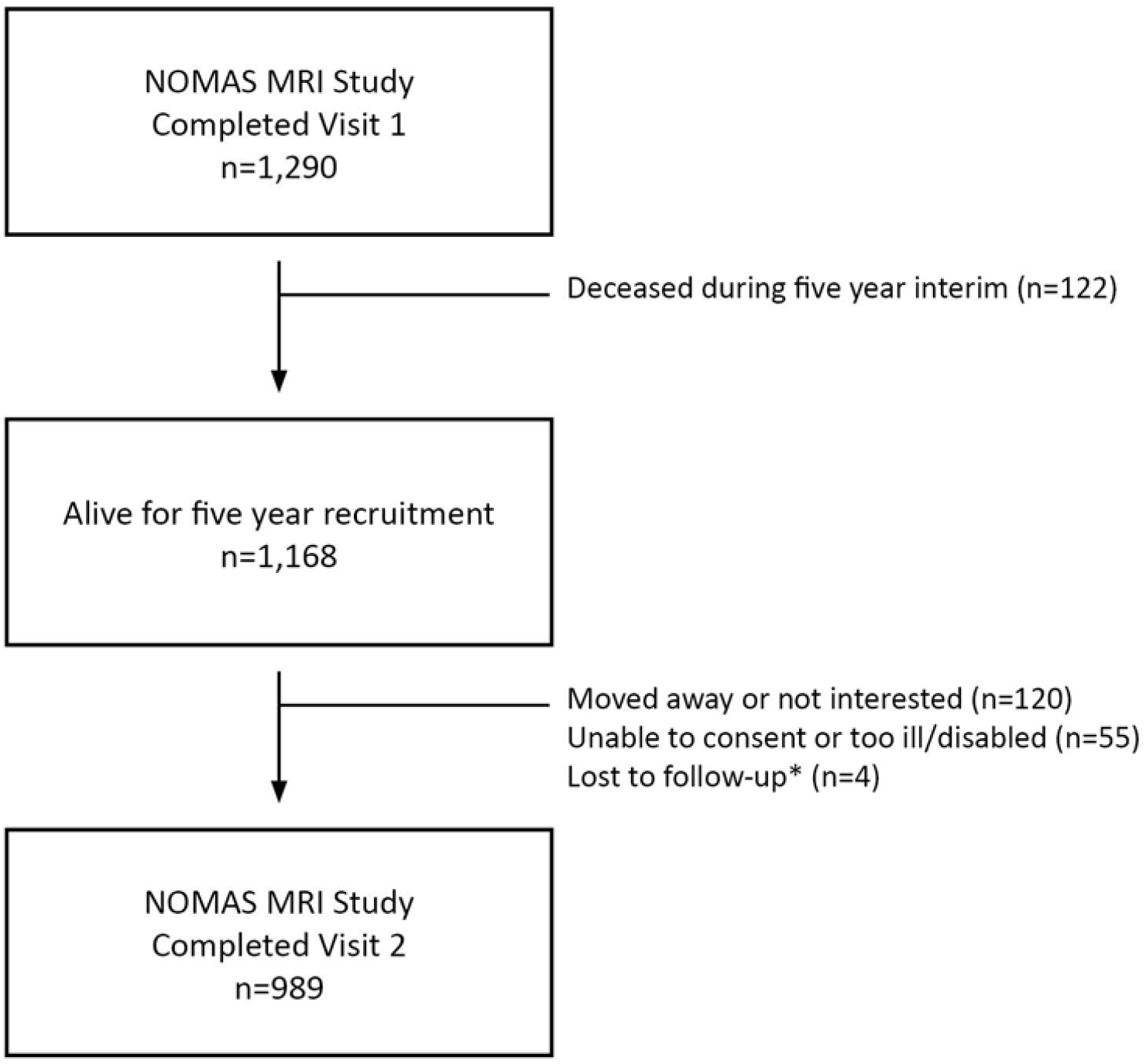
Flow chart showing NOMAS MRI sample participation in first and second in-person assessments. * Loss to follow-up may be temporary as attempts to recontact continue.

**Table 1:** Participant characteristics.

|  | **Total (N=989)** | **Hispanic (N=672)** | **Black (N=162)** | **White (N=134)** | **p-value\*** |
| --- | --- | --- | --- | --- | --- |
| Age, years | 69 ± 9 | 68 ± 8 | 72 ± 9 | 72 ± 9 | <.0001 |
| Years of Education | 10 ± 5 | 8 ± 5 | 13 ± 3 | 15 ± 3 | <.0001 |
| Health Insurance Status |  |  |  |  |  |
| Medicaid or no insurance (%) | 484 (49) | 434 (65) | 35 (22) | 12 (9) | <.0001 |
| Medicare or private insurance | 505 (51) | 238 (35) | 127 (78) | 122 (91) |  |
| Women | 610 (62) | 425 (63) | 111 (69) | 67 (50) | 0.002 |
| Current Smoker | 155 (16) | 97 (14) | 39 (24) | 18 (13) | 0.003 |
| Moderate or Heavy Physical Activity | 97 (10) | 45 (7) | 19 (12) | 28 (21) | <.0001 |
| Diabetes mellitus | 267 (27) | 207 (31) | 42 (26) | 15 (11) | <.0001 |
| Systolic BP, mm Hg | 136 ± 17 | 136 ± 17 | 139 ± 18 | 132 ± 17 | 0.0003 |
| Diastolic BP, mm Hg | 79 ± 9 | 79 ± 9 | 80 ± 10 | 76 ± 10 | 0.0032 |
| Anti-hypertensive use | 590 (60) | 414 (62) | 109 (67) | 60 (45) | <.0001 |
| HDL, mg/dL | 53 ± 17 | 50 ± 14 | 61 ± 20 | 59 ± 20 | <0.0001 |
| LDL, mg/dL | 116 ± 36 | 118 ± 36 | 111 ± 35 | 114 ± 35 | 0.314 |
| Waist Hip Ratio | 0.9 (0.1) | 0.9 (0.1) | 0.9 (0.1) | 0.9 (0.1) | 0.016 |
| Literacy/vocabulary |  |  |  |  |  |
| WRAT and WAT, Z-score | 0.01 ± 1.0 | -0.01 ± 1.0 | -0.25 ± 1.0 | 0.410 ± 0.8 | <.0001 |
| Peabody, Z-score | 0.05 ± 1.0 | 0.02 ± 1.0 | -0.19 ± 1.0 | 0.478 ± 0.8 | <.0001 |
| Percent WMHV** | 0.5 ± 0.7 | 0.5 ± 0.7 | 0.9 ± 1.0 | 0.6 ± 0.8 | 0.0007 |
| TCB *** | 0.73 ± 0.04 | 0.74 ± 0.04 | 0.71 ± 0.04 | 0.71 ± 0.05 | <.0001 |

\* p-values for age adjusted global comparison among 3 race-ethnicity groups (2 degrees of freedom)
\*\* Percent Total Cranial Volume adjusted WMHV
\*\*\* Total Cranial Volume adjusted (proportion)

We ascertained the cognitive status of 989 NOMAS participants who attended visit 2. One hundred ninety-nine participants were adjudicated as having MCI (20%), 45 as having probable dementia (5%), 8 as Other or Unable to Classify (<1%), and 737 participants were adjudicated or identified by algorithm as having no cognitive impairment (74%). The breakdown of cognitive status by race/ethnicity is shown in Table 2. The ordinal odds of MCI or probable dementia was significantly greater for Black and Hispanic participants as a group compared to Whites adjusting for sociodemographic variables (LRT 2df, p<0.0001; Table 3). Table 3 shows that each race/ethnicity had significantly greater odds of MCI or probable dementia than Whites (two-fold for Blacks and four-fold for Hispanics). Adjusting further for vascular risk factors in model 2, the association across race/ethnicity remained significant (LRT 2df, p<0.0001), but the effect was slightly attenuated by about 6% for Black participants and was unchanged for Hispanics (Table 3). No individual risk factor was significantly associated with the odds of MCI or probable dementia in this model (data not shown). When we adjusted for sociodemographic variables and brain imaging markers in model 3, the odds of MCI or probable dementia remained significantly greater for Black and Hispanic participants than Whites (LRT 2df, p<0.0001), but the association was attenuated by almost 25% for Blacks compared to model 1 (OR=1.7, 95% CI=0.9 to 3.32) and again did not change for Hispanics (OR=4.2, 95% CI=2.0 to 8.8). In this model, each unit greater log-WMHV burden was associated with 1.3 times greater odds of MCI or probable dementia (OR=1.3, 95% CI=1.1 to 1.6), but neither smaller BPF nor SBI status was significantly associated with odds of MCI or probable dementia in model 3. There was no significant interaction between Black race and log-WMHV (p for interaction 0.473). Restricting the primary analyses to NOMAS participants with normal MMSE scores (27 or greater) at visit 1, we found similar results (Supplemental Table 2).

**Table 2:** Mild cognitive impairment and probable dementia status by race/ethnicity at visit 2.

| **Cognitive status** | **Total (N=989)** | **Hispanic (N=672)** | **Black (N=162)** | **White (N=134)** | **P-value** |
| --- | --- | --- | --- | --- | --- |
| Normal, N (%)<sup>*</sup> | 737 (74) | 488 (74) | 116 (72) | 114 (86) | <.0001<sup>**</sup> |
| MCI | 199 (20) | 141 (20) | 40 (25) | 16 (12) |  |
| Probable dementia | 45 (5) | 36 (5) | 6 (3) | 3 (2) |  |

<sup>\*</sup> Rounded to whole numbers
<sup>\*\*</sup>Chi-Squared with 4 degrees of freedom

**Table 3:** Multivariable model of race/ethnicity and MCI/probable dementia outcomes.

| Parameter |  | OR | 95% CI of OR |  | p-value* | p-value† |
| --- | --- | --- | --- | --- | --- | --- |
| Model 1 |  |  |  |  |  |  |
| Black | vs. White | 1.9 | 1 | 3.7 | 0.048 | <0.0001 |
| Hispanic vs. White |  | 4.5 | 2.3 | 8.9 | <0.0001 |  |
| Hispanic vs. Black |  | 2.3 | 1.3 | 4 | 0.003 |  |
| Model 2 |  |  |  |  |  |  |
| Black | vs. White | 1.8 | 0.9 | 3.6 | 0.213 | <0.0001 |
| Hispanic vs. White |  | 4.5 | 2.2 | 9.2 | 0.0002 |  |
| Hispanic vs. Black |  | 2.6 | 1.4 | 4.9 | 0.003 |  |
| Model 3 |  |  |  |  |  |  |
| Black | vs. White | 1.7 | 0.9 | 3.3 | 0.113 | <0.0001 |
| Hispanic vs. White |  | 4.3 | 2.2 | 8.5 | <0.0001 |  |
| Hispanic vs. Black |  | 2.5 | 1.4 | 4.4 | 0.001 |  |

\* p-value for pairwise comparison
† p-value for Likelihood Ratio Test with 2 degrees of freedom
Model 1 adjusted for age, education, medical insurance status, literacy.
Model 2 adjusted for variables in model 1 and smoking, leisure time physical activity, diabetes mellitus, systolic and diastolic blood pressure, antihypertensive use, low- and high-density lipoprotein levels, waist hip ratio, and reported ethanol use.
Model 3 adjusted for variables in model 1 and brain parenchymal fraction, subclinical infarct status, and log white matter hyperintensity volume.

## DISCUSSION

This analysis of MCI and dementia prevalence in NOMAS represents the first estimate in the sample since expert adjudication of cognitive status was initiated in 2016. We found prevalence rates of MCI and dementia similar to a number of other cohorts that have examined Hispanic, Black, and White people.^29-31^ For dementia the relatively low prevalence of 5% is similar to other cohorts that included people below the age of 65, and studies have generally reported that the proportion of people with dementia rises dramatically with age.^4^ We found marked race/ethnic differences in this study. Black participants were twice as likely as White participants, and Hispanic participants more than four times as likely than White participants, and twice as likely as Black participants, to have MCI or dementia adjusting for sociodemographic factors.

Racial and ethnic disparities in dementia risk have been known for many years and are well supported by studies in different groups, including in a sample of Medicare participants in Northern Manhattan.^32^ However, more recent data reported temporal trends showing sharp reductions in dementia rates across race/ethnic groups with greater declines for non-Hispanic Blacks and Whites.^33^ The biracial Atherosclerosis Risk in Communities (ARIC) Neurocognitive Study also found a greater prevalence of dementia, but not MCI, in Blacks compared to Whites.^29^ Few studies have included Hispanics to be able to compare MCI and dementia rates across racial and ethnic groups.^4^ Continued follow-up in NOMAS will provide incidence data in a sample that included people as young as 50 at the time of the first evaluation of MCI and dementia status (visit 1).

Given the importance of midlife as opposed to late life vascular risk factors in the development of VCID, opportunities to study these relationships in diverse cohorts are needed. Especially those where participants live in the same community, allowing comparison across groups without confounding introduced by different environments and heterogeneity in other local factors. The greater burden of some vascular risk factors that has been found in studies that have included Blacks or Hispanics might be expected to explain some of these disparities. Interestingly, in this cross-sectional analysis, adjusting for potentially modifiable vascular risk factors did not explain much of the variance in the association of race/ethnicity with MCI and dementia status, although the association for Blacks was attenuated slightly. Not all studies have confirmed link with vascular risk factor exposures. In the ARIC study, Blacks were at elevated risk of dementia compared to Whites but midlife risk factor burden did not play a key role.^3^ Similarly, in the Health, Aging, and Body Composition study, Blacks were at elevated risk of dementia compared to Whites and adjusting for vascular and other comorbidities did not attenuate this effect.^34^ Among Black and Caribbean Hispanic Medicare recipients in Northern Manhattan, diabetes contributed significantly to MCI and dementia risk.^35, 36^ Diabetes was also a risk factor for MCI in the Hispanic Communities Health Study, which includes Caribbean Hispanic participants such as in NOMAS and other groups.^37^ In NOMAS, we did not find that diabetes was a significant contributor to the odds of MCI or dementia prevalence, but continued follow-up is needed. Midlife hypertension has also been associated with MCI and dementia risk in diverse cohorts, but the risk for dementia was greater for White than Black participants in a biracial study.^38, 39^ There are a number of reasons why midlife risk factors would not explain later cognitive outcomes, including variability in duration of exposure and competing risk of death.

Brain imaging studies can help clarify disparities in the downstream effects of exposure to behavioral and vascular risk factors as well as capture evidence of neurodegeneration. White matter lesion burden has been associated with conversion from normal to MCI and rate of decline from MCI to dementia.^40, 41^ This is of particular interest because WMH are related to cerebral small vessel disease, providing an opportunity for prevention through risk factor control especially hypertension.^42^ Both Black and Hispanic participants have been found to have a greater burden of WMH in several studies, and WMH burden was related to ideal cardiovascular health in NOMAS.^43, 44^ In a study among Mexican Americans both hippocampal volume and WMH burden were independent predictors of dementia.^45^ In keeping with this, the current analysis found that WMH burden explained the disparity in odds of MCI and dementia between Black and non-Hispanic Whites. However, sociodemographic variables, vascular risk factors, and brain imaging markers did not explain the disparity between Hispanic and White participants. The reason for this is not certain but degree of risk factor control and length of exposure are not captured by risk factor adjustment and could be of explanatory value.

Strengths of this study include the well-phenotyped diverse cohort that includes behavioral and risk factor data as well as imaging markers of brain integrity. Survival of participants to participate in the MRI sub-cohort yielded a younger, healthier, sample with better cognitive performance than the original population-based NOMAS sample. Given the likelihood of differential dropout due to mortality leading up to the waves of cognitive assessments it is likely that some bias was introduced. We did not conduct analyses of the competing risk of mortality for this study. Prospective data collection is underway and will allow the current prevalence estimates to establish a baseline for incidence studies where competing risk models will be of value. As with all cross-sectional observational studies, unmeasured confounding of risk factors for cognitive and brain health are another potential source of bias.

In conclusion, data from this racially and ethnically diverse cohort study show that Black and Hispanic people were more likely than Whites to have MCI or dementia at the second visit even if they had no more than mild cognitive problems at the first. These disparities in MCI and dementia prevalence were independent of sociodemographic and vascular risk factors, but WMH burden to a large extent explained the disparity between Black and White participants. Prospective data from NOMAS will help clarify these findings in the future.

## Data Availability

Data for this analysis are available upon reasonable request.

## ACKNOWLEDGEMENTS

This study was supported by the National Institute of Neurological Disorders and Stroke (grant NS 29993)

## Supplemental Material

**Supplemental Table 1:** Comparison of participants who did or did not return for visit 2.

| Variable | Returned for Visit 2 |  | p-value |
| --- | --- | --- | --- |
|  | Yes (n=989) | No (n=301) |  |
| Global cognitive domain Z-score at visit 1 (SD) | 0.1 (0.7) | -0.3 (0.8) | <0.0001 |
| Age, visit 1 | 69 (9) | 75 (9) | <0.0001 |
| Education, years | 10 (5) | 10 (5) | 0.905 |
| Race/ethnicity* |  |  |  |
| White | 134 (14) | 57 (19) | 0.007 |
| Black | 162 (16) | 61 (20) |  |
| Hispanic | 672 (68) | 175 (58) |  |
| Sex |  |  |  |
| Women | 610 (62) | 170 (57) | 0.106 |
| Men | 379 (38) | 131 (44) |  |
| Medical insurance status |  |  |  |
| Medicare or private insurance | 505 (51) | 172 (57) | 0.064 |
| Medicaid or no insurance | 484 (49) | 129 (43) |  |
| Literacy Z-scores |  |  |  |
| WRAT/WAT | 0.01 (0.9) | -0.05 (1.04) | 0.387 |
| Vocabulary | 0.05 (1.0) | -0.16 (1.0) | 0.003 |
| Current smoker | 155 (16) | 48 (16) | 0.909 |
| Moderate or Heavy Physical Activity | 97 (10) | 34 (11) | 0.464 |
| Diabetes mellitus | 267 (27) | 86 (29) | 0.592 |
| Systolic BP, mm Hg | 136 (17) | 139 (18) | 0.004 |
| Diastolic BP, mm Hg | 79 (9) | 76 (10) | 0.0005 |
| HDL, mg/dL | 53 (17) | 54.9 (17) | 0.079 |
| LDL, mg/dL | 116 (36) | 114.2 (34) | 0.442 |

\* Column percentages sum to 98 because data for Other races are not included.

**Supplemental Table 2:**
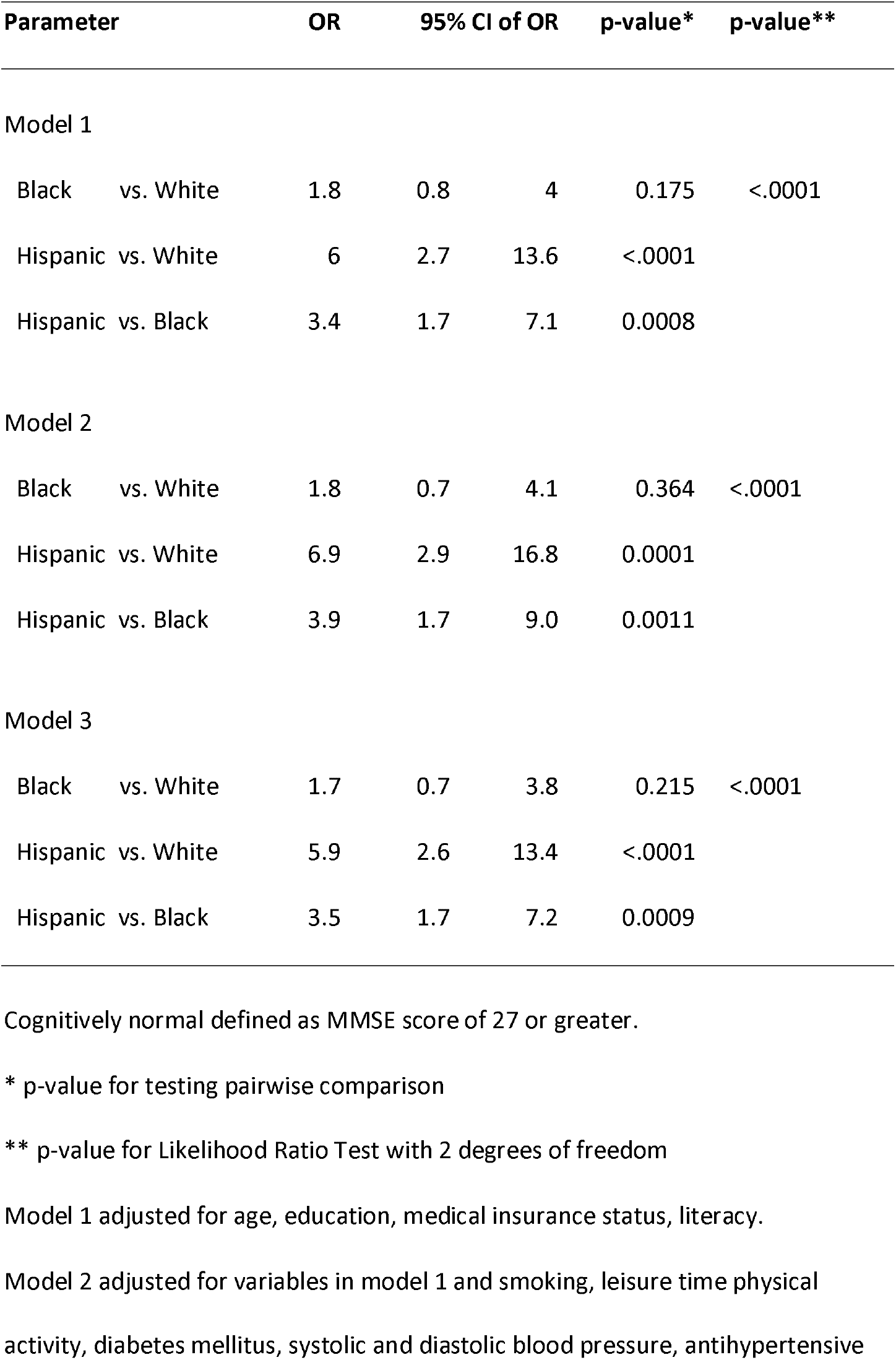
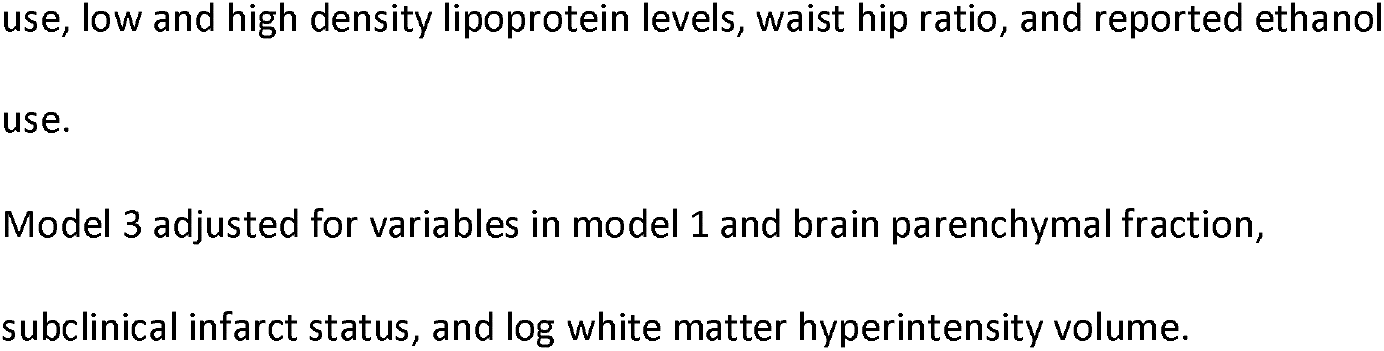
Multivariable model of race/ethnicity and MCI/probable dementia outcomes among participants that were cognitively normal at visit 1

